# Genetic and epigenetic regulation of *SLC6A4* shapes vulnerability to cognitive decline and depressive tendency in later life

**DOI:** 10.64898/2026.03.26.26349121

**Authors:** Yutaro Yanagida, Yutaka Nakachi, Naoto Kajitani, Satoshi Kikkawa, Tempei Ikegame, Shinsuke Koike, Norihide Maikusa, Naohiro Okada, Izumi Naka, Jun Ohashi, Hiroko Sugawara, Kazuhiro Yoshiura, Ayaka Fujii, Emi Kiyota, Risa Watanabe, Yui Murata, Yasuyuki Taki, Yasuko Tatewaki, Benjamin Thyreau, Yuji Takano, Manabu Makinodan, Norio Sakai, Tomohisa Ishikawa, Yosuke Hidaka, Mamoru Hashimoto, Yoshihiko Furuta, Tomoyuki Ohara, Toshiharu Ninomiya, Kiyoto Kasai, Minoru Takebayashi, Miki Bundo, Kazuya Iwamoto, The Japan Prospective Studies Collaboration for Aging and Dementia (JPSC-AD) study group

## Abstract

Age-related cognitive decline and depressive symptoms are prevalent in later life, yet the genetic determinants of vulnerability remain unclear. Here, we investigated how genetic and epigenetic regulation of the serotonin transporter gene *SLC6A4* contributes to susceptibility to these age-related conditions in later life. In community-dwelling older adults in Japan (*N* = 1,317), functional stratification of the serotonin transporter-linked polymorphic region (5-HTTLPR) revealed that participants with low-activity genotypes showed a robust co-occurrence of cognitive decline and depressive symptoms, whereas this comorbid pattern was not observed in those with the high-activity genotype. The genotype-dependent co-occurrence was consistently replicated across seven independent population-based cohorts (total *N* = 7,889). DNA methylation at a functional promoter CpG site increased with age and partially mediated age-related cognitive decline specifically among low-activity genotypes. In contrast, the high-activity genotype was associated with relative resistance to these functional declines, partly mediated by a protective effect on hippocampal volume during aging. Notably, genotype-dependent effects on hippocampal volume were absent in adolescence, indicating that the influence of *SLC6A4* emerges in an aging-specific manner. Together, these findings identify *SLC6A4* promoter activity as a key genetic factor modulating vulnerability and resilience in later life.

## Introduction

Serotonin (5-hydroxytryptamine, 5-HT) modulates a wide-range of physiological and psychological functions, including cognitive processes and mood regulation (1–4). Across the adult lifespan, serotonergic signaling gradually declines, a process implicated in both cognitive impairment and depressive symptoms observed in later life (5,6). The hippocampus, a region critical for memory and emotional processing (7), receives dense serotonergic projections essential for maintaining its structural integrity and neuroplasticity (1,3,4). During aging, alterations in these pathways are associated with progressive hippocampal volume reduction, which has been linked to the emergence of cognitive and affective decline (2,5,8–13). Several studies have suggested that lateralized hippocampal functions, including those involving the right hippocampus, may contribute to emotional regulation and episodic memory (7,14,15), This raises the possibility that structural variation in hippocampus may serve as a key neurobiological mediator linking the serotonergic system to these clinical outcomes.

Serotonin transporter (5-HTT), encoded by *SLC6A4*, plays a critical role in serotonergic neurotransmission by regulating synaptic 5-HT availability through reuptake (3,4). Transcriptional regulation of *SLC6A4* is influenced by genetic variation, most notably the 5-HTT-linked polymorphic region (5-HTTLPR), a functional variable numbers of tandem repeats located in the promoter region. The 5-HTTLPR has traditionally been classified into short (S) and long (L) alleles based on repeat length, with the L allele showing the higher promoter activity (16–18). This polymorphism has been extensively investigated in relation to stress sensitivity and vulnerability to depression and related psychopathology, with early works demonstrating moderation of life stress effects by genotype (19–21). Although numerous association studies have reported links between the S allele and depression or other psychiatric disorders, simple genotype-phenotype associations were not supported in recent large-scale studies (22,23).

Accumulating evidence indicates that the functional complexity of the 5-HTTLPR locus needs to be considered beyond the binary S/L classification. More than eight S-type alleles, eight L-type, and several atypical variants have been identified, with substantial differences in allele composition across populations (24). In the Caucasian and European populations, the major allele is L_16a_ (allele frequency approximately 51%), followed by the S (40%) and L_16d_ (9%) (25). By contrast, in the Japanese population, the S allele predominates (78.8%), followed by three L alleles (L_16a_, L_16c_ and L_16d_), each with similar allele frequencies ranging about 5-6 % (18,24). Importantly, only the L_16a_ allele showed high promoter activity, whereas other L alleles (L_16c_ and L_16d_) showed low promoter activity comparable to the S allele. Although several studies have attempted to account for three alleles (L_16a_, S and L_16d_), classification based solely on repeat length remains in sufficient, and a functionally based approach, such as promoter activity–based classification, is useful for capturing the diversity of the 5-HTTLPR (23,25,26).

Epigenetic regulation adds an additional layer of complexity. DNA methylation at the promoter CpG island and its neighboring region of *SLC6A4* modulates transcriptional activity, and certain CpG sites have been shown to exert strong functional effects (27). Previous studies identified two such CpG sites, referred to as CpG3 (hg19 coordinates, chr17:30,235,246–30,235,247) and CpG4 (chr17:30,235,271–30,235,272), which have been strongly implicated in psychiatric disorders and pharmacological treatment response, with CpG3 hypermethylation leading to near-complete loss of promoter activity *in vitro* (26,28,29).

As outlined above, both genetic and epigenetic regulation of *SLC6A4* have been extensively investigated in relation to stress sensitivity, depression, and other psychiatric conditions with onset in early or mid-adulthood. However, their relevance to age-related cognitive and affective changes during normal aging remains largely unclear, except for a few pioneering studies (30–33). These gaps underscore the need to re-evaluate the role of *SLC6A4* across the life course, as genetic and epigenetic influences may shift with aging-related changes in serotonergic function, brain structure, and epigenetic regulation. In addition, large-scale studies in East Asian populations, where low-activity 5-HTTLPR alleles are prevalent, remain limited, despite their potential to reveal biological patterns not captured in predominantly European cohorts.

In this study, we investigated how genetic and epigenetic regulation of *SLC6A4* relates to cognitive function and depressive tendency in later life. We first examined promoter activity-dependent effects of *SLC6A4* in community-dwelling older adults using the Arao cohort, a part of the Japan Prospective Studies Collaboration for Aging and Dementia (JPSC-AD) (34). We then validated key findings across multiple independent cohorts within JPSC-AD. To determine whether the findings represent stable features established earlier in life or instead emerge preferentially with aging, we incorporated a population-based adolescent cohort with available genetic and neuroimaging data (35). The age-contrast design identified aging-specific effects of *SLC6A4*, providing a life-course perspective on vulnerability and resilience in later life.

## Results

### Functional stratification of *SLC6A4* in older adults

After participant selection and quality control, the final analytic sample comprised 1,317 community-dwelling older adults aged 65-99 years (mean age, 74.0 ± 6.5 years), including 507 males (38.5%), from the Arao cohort of the JPSC-AD (34) (**Fig. 1a**). Given the considerable heterogeneity of 5-HTTLPR alleles, analyses were restricted to the four major genotypes, which together accounted for 91.8% of the cohort (24) (**Fig. 1b and Supplementary Table 1**). These genotypes comprised S_14a_/S_14a_, L_16c_/S_14a_, L_16d (16g)_/S_14a_ and L_16a_/S_14a_, and were classified based on promoter activity, with the first three designated as low-activity (*N* = 1,054) and L_16a_/S_14a_ as high-activity (*N* = 154). Because all genotypes contained the S_14a_ allele in either homozygous or heterozygous form, we hereafter refer to both the alleles and the corresponding genotypes simply as 14a, 16c, 16d, and 16a, respectively.

**Fig. 1.**
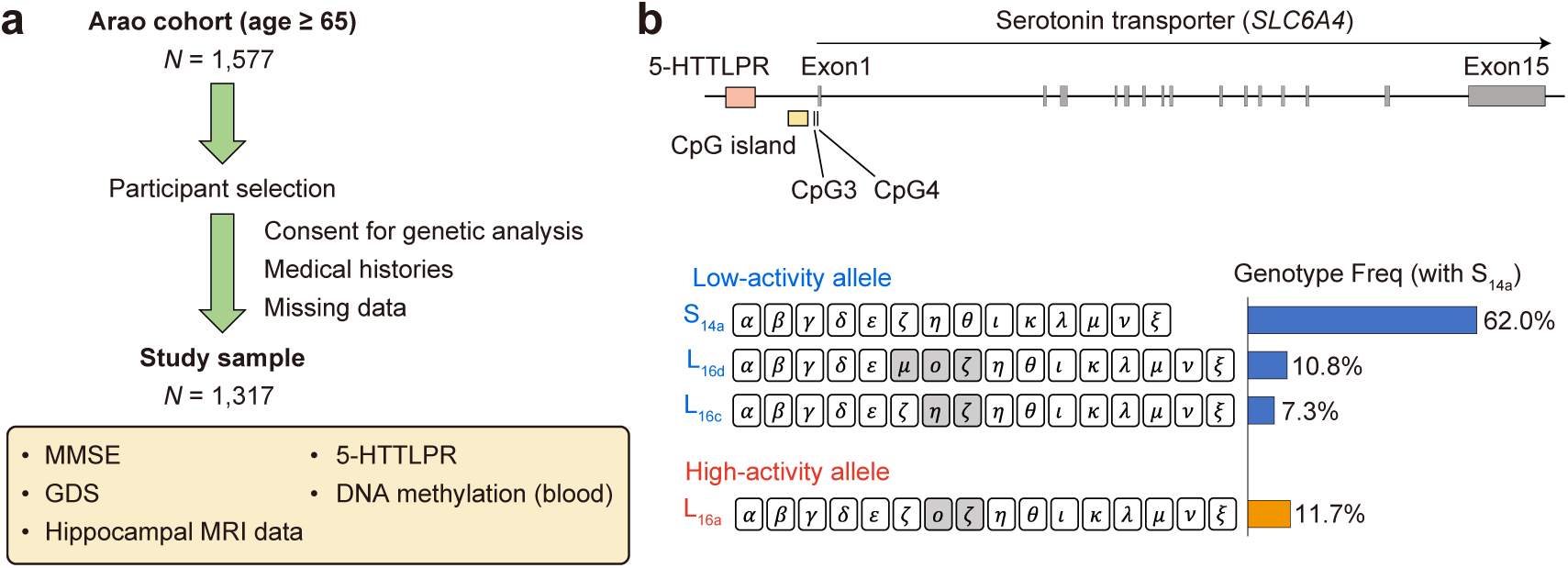
Study design and functional stratification of *SLC6A4* in the Arao cohort. **a,** Participant selection from the Arao cohort. Among 1,577 community-dwelling individuals aged ≥65 years, participants were included based on consent for genetic analysis, availability of medical history, and completeness of data, yielding a final analytic sample of 1,317 participants. Data on cognitive function (MMSE), depressive tendency (GDS), hippocampal MRI, 5-HTTLPR genotype (24), and DNA methylation of the *SLC6A4* promoter were analyzed. **b,** Genomic structure of the serotonin transporter gene *SLC6A4* and functional stratification of the 5-HTTLPR. The locations of CpG3 and CpG4 (hg19 coordinates; CpG3: chr17:30,235,246–30,235,247 and CpG4: chr17:30,235,271–30,235,272) near the promoter CpG island are shown, and their DNA methylation levels were measured. The four major 5-HTTLPR alleles, accounting for 97.8% of the cohort (**Supplementary Table 1**), are shown. The 5-HTTLPR consists of a repetitive array of distinct 20-23 bp units, denoted by Greek letters; differences from the 14a allele are indicated by shaded boxes. Genotype frequencies among individuals carrying the S_14a_ allele are shown on the right. Low-activity and high-activity genotypes are highlighted in blue and red, respectively.

### Promoter activity-dependent association between cognitive decline and depressive tendency

Cognitive function and depressive tendency were quantified as continuous measures using the Mini-Mental State Examination (MMSE) (36,37) and the Geriatric Depression Scale (GDS) (38–41), respectively. The mean and SD of the MMSE and GDS scores were 27.1 ± 3.0 and 2.3 ± 2.3, respectively, capturing inter-individual variability largely within the range of normal aging (**Supplementary Table 2**). Focusing on the influence of age and sex, MMSE scores showed a significant negative correlation with age (Pearson’s R = −0.39, *P*-value < 0.01), whereas GDS scores exhibited a significant positive correlation (R = 0.09, *P*-value < 0.01). No significant differences in these clinical scores were observed between male and female groups (Mann-Whitney *U* test, *P*-value > 0.05).

We first examined the effects of 5-HTTLPR on MMSE and GDS scores. No significant differences in MMSE or GDS scores were observed between the high- and low-activity genotypes (Mann-Whitney *U* test, *P*-value > 0.05) or across four major genotypes (Steel’s multiple comparison test, *P*-value > 0.05) (**Fig. 2ab and Supplementary Table 2**). However, MMSE scores inversely correlated with GDS scores, indicating an association between cognitive decline and depressive tendency, in participants with low-activity genotypes (Pearson’s R = −0.19, *P*-value = 9.97 × 10^-10^) (**Fig. 2c**). This association showed a consistent trend across the low-activity genotypes but was not detected in the high-activity genotype (R = −0.02, *P*-value = 0.77). Sensitivity analysis using logistic regression, adjusted for age and sex, showed that MMSE-defined cognitive impairment was significantly associated with a higher risk of depressive tendency in participants with the low-activity genotype (odds ratio [OR] = 2.02; 95% CI, 1.23 to 3.25; *P*-value = 0.004), but not in those with the high-activity genotype (OR = 0.58; 95% CI, 0.08 to 2.59; *P*-value = 0.52).

**Fig. 2.**
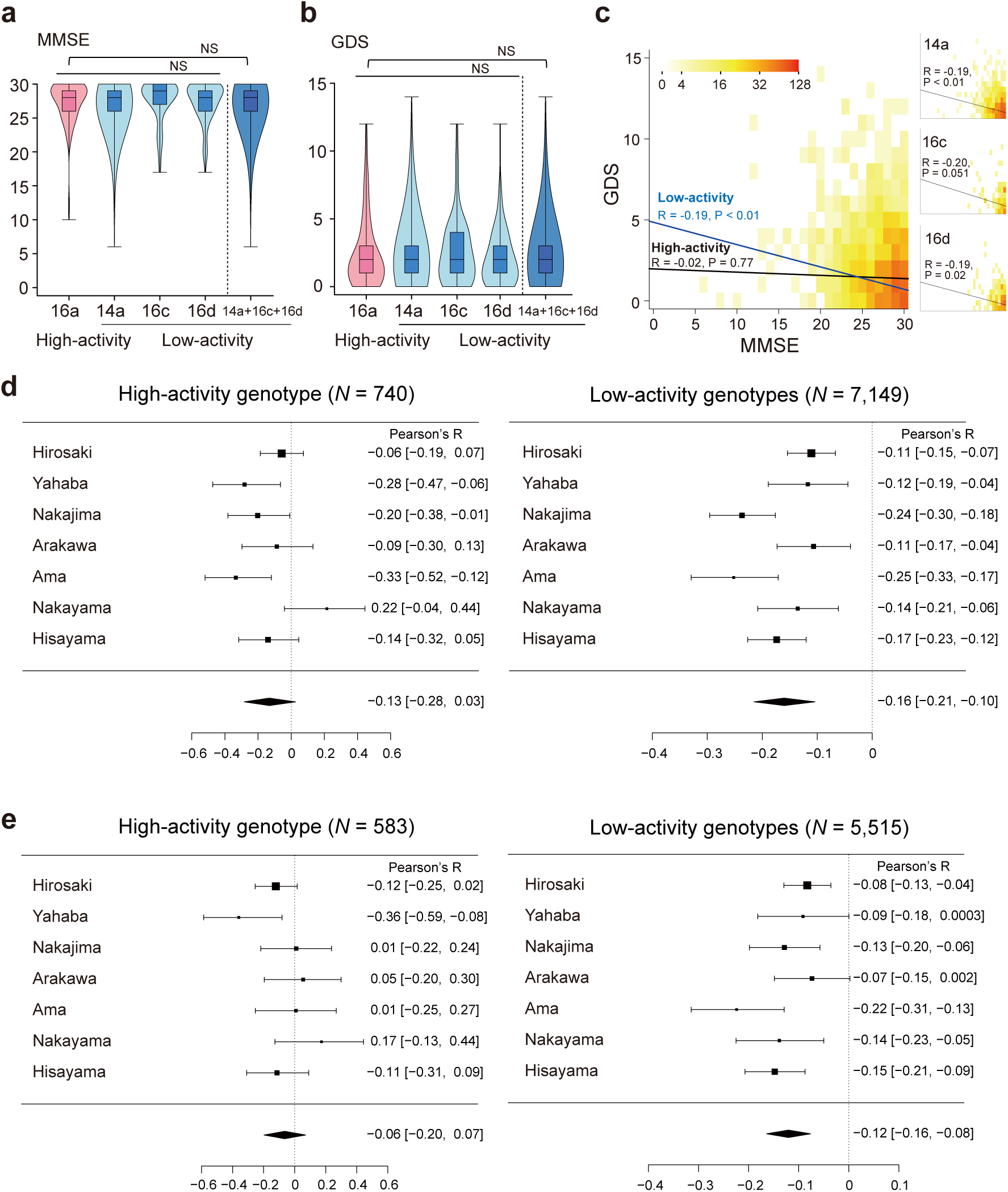
Promoter activity–dependent association between cognitive function and depressive symptoms. **a, b,** Distributions of Mini-Mental State Examination (MMSE) scores (a) and Geriatric Depression Scale (GDS) scores (b) across 5-HTTLPR genotypes stratified by promoter activity. Decreasing of the MMSE score indicates cognitive decline and increasing of the GDS score indicates depressive tendency. Genotypes were grouped into high-activity (16a) and low-activity (14a, 16c, 16d) categories. Violin plots show the full distribution with embedded box plots indicating the median and interquartile range. No significant differences in MMSE or GDS scores were observed across genotypes (NS, not significant in Steel’s multiple comparison test or Mann-Whitney *U* test). **c,** Density plot illustrating the relationship between MMSE and GDS scores in the Arao cohort. An inverse correlation between cognitive decline and depressive tendency was observed in participants with low-activity genotypes (blue line), whereas no such association was detected in participants with the high-activity genotype (black line). Insets show genotype-specific correlations for each low-activity genotype. **d, e,** Forest plots summarizing cohort-specific correlations between MMSE and GDS scores stratified by promoter activity across JPSC-AD cohorts, for all participants (d) and participants without dementia or depression (e). Left panel shows participants with the high-activity genotype, and right panel shows participants with low-activity genotypes. Points indicate Pearson’s correlation coefficients for each cohort with 95% confidence intervals; diamonds represent pooled estimates from meta-analysis. A consistent inverse association was observed only in the low-activity genotype group.

To confirm the association in low-activity genotypes, we further analyzed data from seven additional cohorts within the JPSC-AD study (**Extended Data Fig. 1a**). We applied the same sample selection criteria used for the Arao cohort, and a total of 7,889 participants were included in the analysis. We then imputed 5-HTTLPR genotypes using a method that accounts for the complexity of the 5-HTTLPR variants (42). The frequencies of major 5-HTTLPR alleles and genotypes were comparable to those observed in the Arao cohort (**Extended Data Fig. 1b and Supplementary Table 3,4**). Using the imputed genotypes, the inverse association between MMSE and GDS scores among participants with low-activity genotypes was consistently replicated across all cohorts (range of Pearson’s R: −0.25 to −0.11). A meta-analysis further supported this finding, yielding a pooled Pearson’s R of −0.16 (95% CI, −0.21 to −0.10) (**Fig. 2d**). In contrast, the association between MMSE and GDS scores was not reproducibly observed among participants with the high-activity genotype (range of Pearson’s R: - 0.33 to 0.22) and was not supported by meta-analysis (pooled Pearson’s R = −0.13; 95% CI, −0.28 to 0.03) (**Fig. 2d**).

### Persistent sex differences and age-related increases in promoter methylation

We measured DNA methylation levels of two CpG sites (CpG3 and CpG4) in the promoter region of *SLC6A4* by pyrosequencing assay (**Fig. 1b and 3a**). Consistent with previous reports (26,43,44), DNA methylation levels were significantly higher in females than in males in older adults (CpG3, *P*-value = 5.45 × 10^-46^; CpG4, *P*-value = 7.62 × 10^-41^, in Mann-Whitney *U* test) (**Fig. 3b and Extended Data Fig. 2a**).

**Fig. 3.**
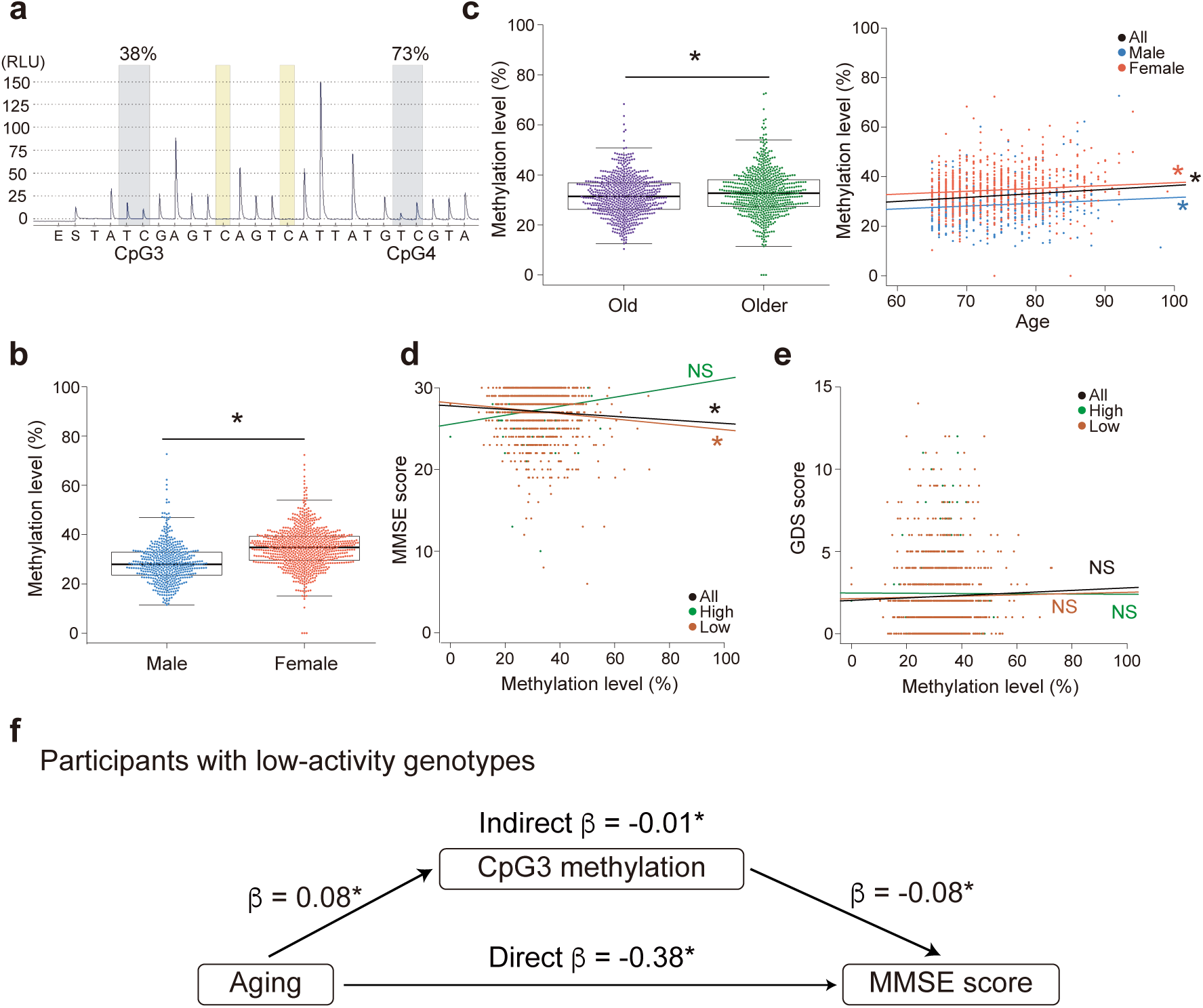
Age- and genotype-dependent effects of *SLC6A4* promoter DNA methylation on cognitive function. **a,** A representative pyrogram. The vertical axis indicates light strength with peaks of nucleotides incorporation (RLU, relative light unit). Blue and yellow squares indicate targeted cytosines for methylation level and bisulfite conversion control, respectively. This pyrogram indicates 38% and 73% methylation at CpG3 and CpG4, respectively. **b,** DNA methylation levels at CpG3 in males (*N* = 507) and females (*N* = 810). **c,** DNA methylation levels at CpG3 increased with age. (*left*) Methylation levels at CpG3 in old (*N* = 671) and older (*N* = 646) participants, divided by the median age of 74. (*right*) Correlation analysis with age. Black, blue, and red colors indicate individual data or regression line of all participants, males, and females, respectively. **d,** Correlation analysis with the MMSE score. Black, green, and brown colors indicate individual data or regression line of all participants, participants with the high-activity genotype (*N* = 154), and those with low-activity genotypes (*N* = 1,054), respectively. **e,** Correlation analysis with the GDS score. **f,** Mediation analysis of CpG3 methylation in participants with low-activity genotypes. A variable of sex was utilized as a covariate. Regression coefficients are shown for each path. Box plots indicate median, interquartile range, and whiskers extending to 1.5× the interquartile range. Asterisks indicate *P*-value < 0.05. NS, not significant.

We further observed that DNA methylation levels were significantly higher in older participants compared with younger participants within the cohort, when stratified by the median age of 74 years (CpG3, *P*-value = 3.32 × 10^-3^ in Mann-Whitney *U* test) (**Fig. 3c**). In addition, DNA methylation levels showed positive correlations with age in the overall sample and when analyzed separately in males and females, independent of 5-HTTLPR genotype (all participants, Pearson’s R = 0.12, *P*-value = 9.01 × 10^-6^; male, R = 0.09, *P*-value = 0.04; female, R = 0.09, *P*-value = 8.1 × 10^-3^) (**Fig. 3c**). Weaker but similar age-related patterns were observed at CpG4 and are summarized in **Extended Data Fig. 2b**.

### Promoter activity-dependent association between CpG3 methylation and cognitive function

DNA methylation levels at CpG3 were negatively correlated with MMSE score (Pearson’s R = −0.06, *P*-value = 0.03) (**Fig. 3d**). This negative correlation was observed exclusively in participants with low-activity genotypes (R = −0.09, *P*-value = 2.37 × 10^-3^), and was not detected in those with the high-activity genotype (R = 0.13, *P*-value = 0.10). Multiple regression analysis confirmed that this association was not confounded by sex or age (β = −0.08, *P*-value = 5.75 × 10^-3^). In contrast, DNA methylation levels at CpG3 were not correlated with GDS score, either in the overall samples (R = 0.03, *P*-value = 0.30) or among participants with low-activity genotypes (R = 0.01, *P*-value = 0.63) (**Fig. 3e**). No significant correlations with either MMSE or GDS scores were observed at CpG4 (**Extended Data Fig. 2cd**).

### CpG3 methylation mediates age-related cognitive decline but not depressive tendency

Given the correlations of CpG3 DNA methylation with age and MMSE score in participants with low-activity genotypes, we next tested whether CpG3 methylation mediated the relationship between aging and the cognitive functions (**Fig. 3f**). Aging showed a significant direct effect on lower MMSE score (β= −0.38, *P*-value < 2.0 × 10^-16^), and CpG3 methylation exerted a significant mediating effect on this association (indirect effect; β = −0.01, *P*-value = 0.03). In contrast, although aging had a significant direct effect of aging on higher GDS score (β = 0.10, *P*-value = 0.03), no significant mediation by CpG3 methylation was detected for depressive tendency (β = 1.33 × 10^-3^, *P*-value = 0.61).

### High-activity genotype has a positive effect on the right hippocampal volume

We performed multiple regression analyses with right and left hippocampal volumes measured by MRI as dependent variables. Age, sex, and intracranial volume (ICV) were included as covariates, and DNA methylation levels or 5-HTTLPR genotypes were examined in separate models as primary explanatory variables. Age, sex, and ICV showed significant effects on bilateral hippocampal volumes (data not shown). DNA methylation levels were not significantly associated with hippocampal volumes (*P*-value > 0.05). In contrast, the high-activity genotype showed a significant positive association with right hippocampal volume (β = 0.07, *P*-value = 0.03) (**Table 1**). This positive effect remained significant when ICV-normalized hippocampal volume was used as the dependent variable (**Supplementary Table 5**).

**Table 1.** Multiple regression analysis of right hippocampal volume. Results of a standardized multiple regression analysis examining the factors associated with right hippocampal volume in older adults (*N* = 1,286). ICV, intracranial volume; SE, standard error; Statistic, *t*-statistic; CI, confidence interval.

| Variable | $\beta$ | SE | Statistic | *P*-value | 95% CI |
| --- | --- | --- | --- | --- | --- |
| Age | -0.50 | 0.02 | -22.35 | < 0.01 | -0.55, -0.46 |
| Sex | 0.10 | 0.03 | 3.61 | < 0.01 | 0.05, 0.16 |
| ICV | 0.33 | 0.03 | 11.31 | < 0.01 | 0.27, 0.39 |
| High activity | 0.07 | 0.03 | 2.16 | 0.03 | 0.01, 0.13 |
| Low activity (14a) | 0.05 | 0.04 | 1.33 | 0.18 | -0.03, 0.13 |
| Low activity (16c) | 0.02 | 0.03 | 0.75 | 0.45 | -0.04, 0.08 |
| Low activity (16d) | 0.03 | 0.03 | 0.95 | 0.34 | -0.03, 0.09 |

To further localize the structural correlates of the association between the high-activity genotype and hippocampal volume, we then performed multiple regression analyses of 21 subfields of the right hippocampus using the available data (*N* = 1,267). The high-activity genotype was nominally significantly associated with larger volumes of the hippocampal tail and the granule cell and molecular layer of the dentate gyrus body (GC.ML.DG.body) (**Extended Data Table 1**). Direct volume comparisons showed differences in the same direction as those identified by multiple regression analyses, with modestly larger right hippocampal and its subfields volumes in participants with the high-activity genotype (**Extended Data Fig. 3**).

### 5-HTTLPR does not affect hippocampal volumes in the youth

To determine whether the effect of the high-activity genotype on right hippocampal volume emerges specifically with aging or is already present earlier in development, we examined this association in the TokyoTEEN Cohort (TTC), a prospective population-based birth cohort study conducted during adolescence (45). Within the TTC, a subset of participants enrolled in the population-neuroscience study (pn-TTC) (35) underwent longitudinal MRI assessments at four developmental stages (ages 11, 13, 15, and 18). We conducted genotyping of 5-HTTLPR and utilized previously reported data on DNA methylation levels and hippocampal volumes from pn-TTC participants (44) (**Supplementary Table 6**). Multiple regression analyses revealed no significant association between the high-activity genotype and right hippocampal volume across any of the four stages, which was further confirmed by linear mixed models analyzing these longitudinal data (*P*-value > 0.05) (**Fig. 4a and Supplementary Table 7**). Furthermore, no significant association was observed between 5-HTTLPR genotype and intra-individual changes in right hippocampal volume, as measured by the ratio of volume at pn-TTC-4 to that at pn-TTC-1 (*P*-value > 0.05). In addition, in the subset with available DNA methylation data, methylation levels at CpG3 and CpG4 showed no significant association with hippocampal volume at any time point (**Fig. 4b**).

**Fig. 4.**
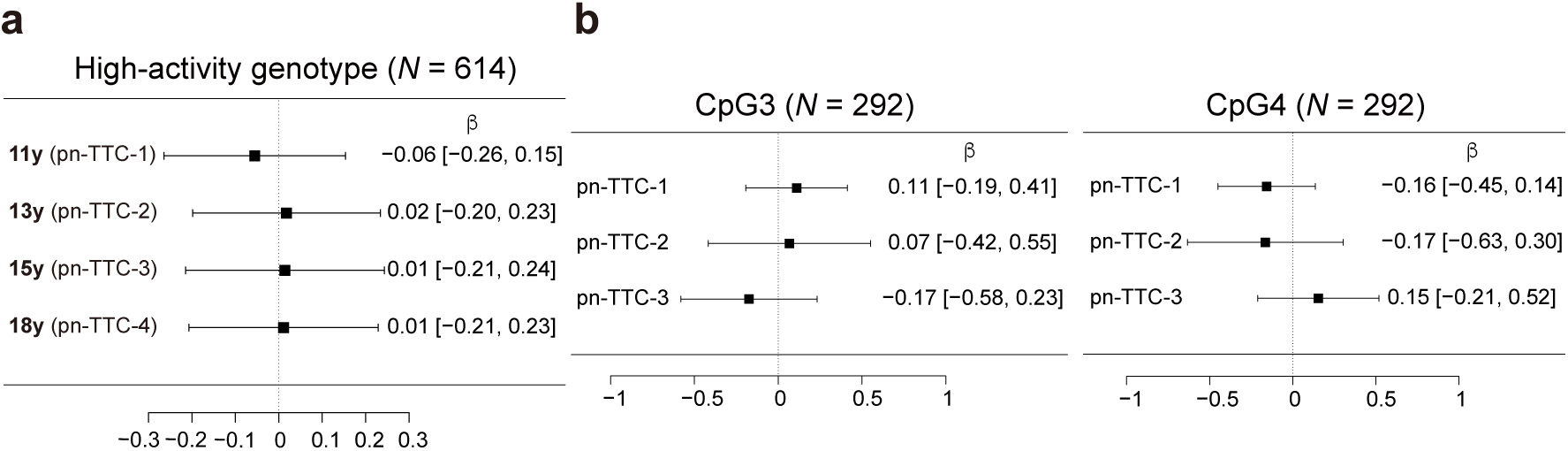
Multiple regression analysis of right hippocampal volume in youth. Forest plots summarizing associations with right hippocampal volume in the longitudinal pn-TTC cohort. **a**, Association between right hippocampal volume and 5-HTTLPR genotype (total *N* = 614). Pn-TTC-1 to −4 represent data from surveys conducted at ages 11, 13, 15, and 18, respectively. **b**, Associations of right hippocampal volume with CpG3 and CpG4 DNA methylation levels (total *N* = 292). Note that DNA methylation data were available for pn-TTC-1 through pn-TTC-3. Points indicate partial regression coefficients for each survey wave with 95% confidence intervals.

### High-activity genotype indirectly affects clinical scores through hippocampal volume

Given that the high-activity genotype showed no direct association with cognitive decline or depressive tendency (**Fig. 2ab**), but was positively associated with right hippocampal volume (**Table 1**), we conducted a mediation analysis to examine whether right hippocampal volume mediates the relationship between this genotype and clinical scores (**Fig. 5a**). Consistent with earlier analyses, no direct effects of the high activity genotype on MMSE or GDS scores were detected. However, right hippocampal volume significantly mediated the association between the high-activity genotype and MMSE (β = 0.02, *P*-value = 0.03). A similar mediating effect was observed for GDS (β = −0.01, *P*-value = 0.03). Subfield-level analyses further supported these findings, with significant or trend-level mediation effects observed in the hippocampal tail and the GC.ML.DG body subfield for both MMSE and GDS (**Extended Data Fig. 4**).

**Fig. 5.**
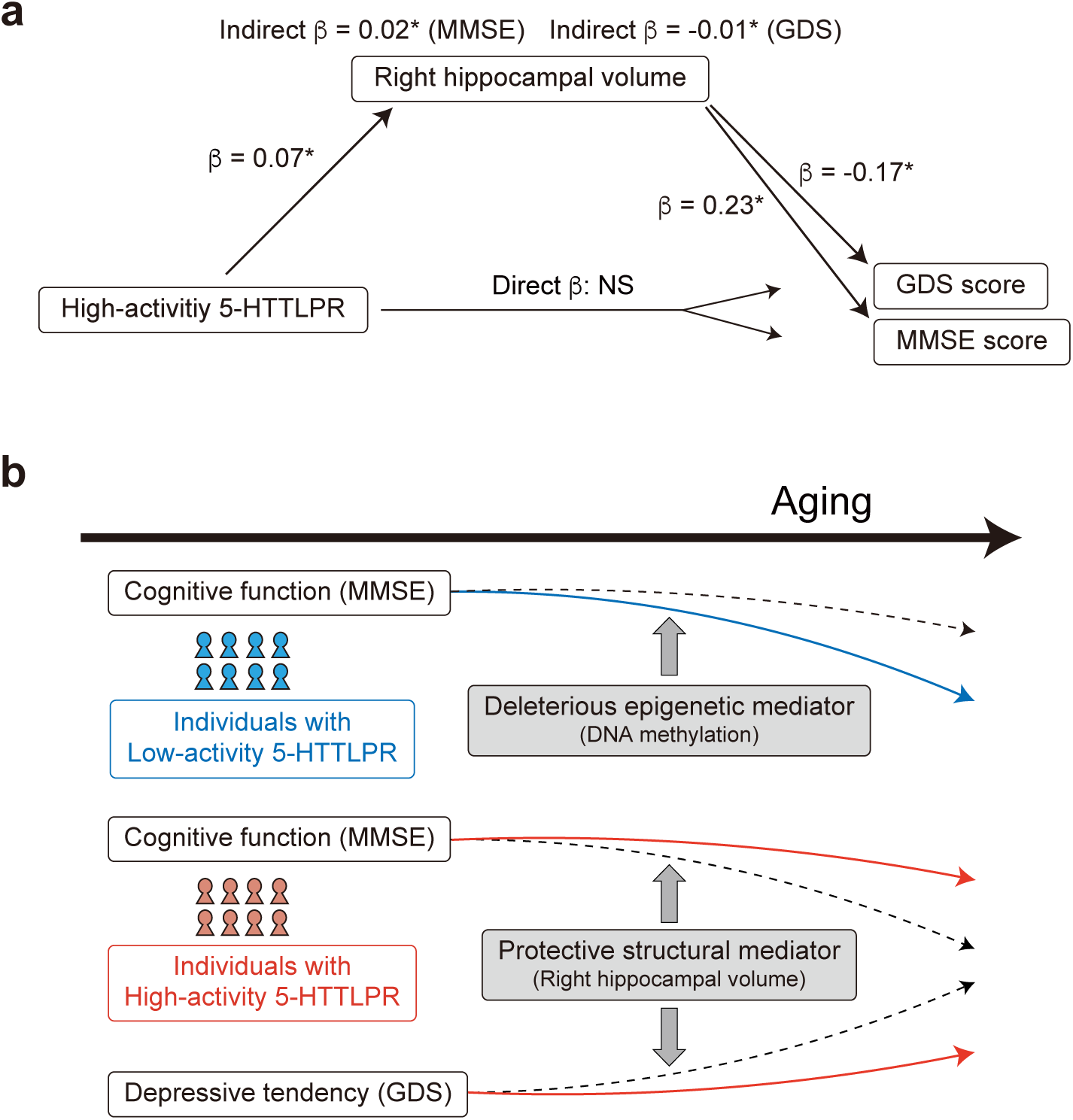
Mediation analysis via right hippocampal volume and graphical summary. **a,** Mediation models of the relationships among the high-activity 5-HTTLPR genotype, right hippocampal volume, and clinical outcomes. Path coefficients (β) are shown for each association. The high-activity genotype was positively associated with right hippocampal volume, which in turn was significantly associated with MMSE and GDS scores. No significant direct effects of the high-activity genotype on MMSE or GDS scores were observed. In contrast, significant indirect effects mediated by right hippocampal volume were detected for both MMSE and GDS, indicating that hippocampal structure mediates the association between 5-HTTLPR promoter activity and cognitive and depressive measures. Asterisks indicate *P*-value < 0.05. NS, not significant. **b**, Graphical summary of the present study, illustrating the associations between 5-HTTLPR genotype and clinical outcomes, as mediated by epigenetic and brain structural factors. Black and red lines represent the divergent trajectories for individuals with low- and high-activity 5-HTTLPR genotypes, respectively.

### Robust replication in cognitively and psychiatrically healthy older adults

Our analyses in the Arao Cohort included individuals diagnosed with dementia (*N* = 36) or depression (*N* = 6) with no overlap between these groups. These conditions may be associated with atypical distributions of clinical scores compared to normally aged participants (37,40). Therefore, we re-examined the analyses after excluding these participants. Following exclusion, the inverse correlation between MMSE and GDS scores remained significant among participants with low-activity genotypes (R = −0.16, *P*-value = 1.48 × 10^-7^) in the Arao cohort. Importantly, these genotype-dependent associations were consistently replicated by meta-analysis of JPSC-AD cohorts when analyses were restricted to participants without dementia or depression, comprising a total of 5,515 participants with low-activity genotypes and 583 with high-activity genotypes (**Fig. 2e**).

We also confirmed that the high-activity genotype retained a significant positive association with right hippocampal volume (β = 0.09, *P*-value = 0.01) and continued to show indirect associations with MMSE (β = 0.01, *P*-value = 0.01) and GDS scores (β = −0.01, *P*-value = 0.01) through hippocampal volume. In contrast, the mediation effect of CpG3 DNA methylation on the relationship between aging and MMSE in participants with low-activity genotypes was no longer statistically significant (β = −4.81 × 10^-3^, *P*-value = 0.09). This attenuation likely reflects the exclusion of participants with lower MMSE scores and is consistent with the possibility that CpG3 methylation is more strongly associated with pathological cognitive decline.

## Discussion

Despite the well-established importance of the serotonergic system in late-life depression and cognitive decline, the role of *SLC6A4* in older adults has remained insufficiently explored. We demonstrate that 5-HTTLPR variation in *SLC6A4* influences cognitive and affective outcomes in later life in a promoter activity–dependent manner. Subjects with low-activity genotypes showed greater vulnerability to both cognitive decline and depressive tendency, with epigenetic modulation further contributing to the severity of cognitive decline. In contrast, participants with the high-activity genotype exhibited relative resistance to these functional declines, partly mediated by a protective effect on hippocampal volume during normal aging (**Fig. 5b**). Notably, these genotype-dependent effects were evident in older adults but were not observed during adolescence, indicating that the functional impact of *SLC6A4* emerges preferentially in the context of aging.

### Genotype-dependent coupling between cognitive decline and depressive symptoms in aging

We observed a significant inverse correlation between MMSE and GDS scores in participants with low-activity 5-HTTLPR genotypes, suggesting that declines in cognitive function and increases in depressive tendency tend to co-occur within this genetically defined subgroup. Late-life depression is frequently accompanied by cognitive impairment, and cognitive decline itself increases vulnerability to depressive symptoms in older adults (46–48). However, direct correlations between MMSE and GDS scores have rarely been demonstrated in cognitively and psychiatrically normal aging populations (49). Our findings extend these observations by indicating that such coupling becomes evident when individuals are stratified by serotonergic genetic background. In participants with low-activity 5-HTTLPR genotypes, reduced transcriptional capacity of *SLC6A4* may lower the threshold at which age-related neural or molecular perturbations concurrently affect cognitive and affective processes. The absence of a comparable association in participants with the high-activity genotype suggests that preserved serotonin transporter function may confer resilience against the convergence of cognitive and affective decline.

### Epigenetic regulation of *SLC6A4* in aging

DNA methylation at CpG3 within the *SLC6A4* promoter was consistently higher in females and increased with age. Sex-associated methylation difference at this locus is established early in development and maintained through adulthood (26,43,44,50), and our results further demonstrate that this difference persists into old age, supporting stability across the life course. Beyond this stable sex effect, promoter methylation showed a modest but significant age-related increase independent of 5-HTTLPR promoter activity and sex, suggesting gradual epigenetic downregulation of *SLC6A4* with aging. In contrast, aging-related CpG3 hypermethylation was not observed in pn-TTC participants (*P*-value > 0.05), indicating that this epigenetic alteration is specific to older adults.

Notably, CpG3 methylation was associated with cognitive decline, but not depressive tendency, specifically in participants with low-activity 5-HTTLPR genotypes. Mediation analysis further indicated that CpG3 methylation partially mediates the association between aging and cognitive decline, but not depressive tendency, in participants with low-activity genotypes, implicating epigenetic regulation of *SLC6A4* as one pathway contributing to age-related cognitive decline. Importantly, this mediation effect was attenuated when participants with dementia were excluded, suggesting that CpG3 methylation may be more strongly associated with pathological cognitive decline rather than with general age-related cognitive decline. Future longitudinal studies will be necessary to clarify the temporal dynamics and causal mechanisms linking *SLC6A4* methylation to cognitive decline in later life.

### Hippocampal structure as a mediator of genetic resilience in aging

We identified the right hippocampus as a brain region through which genetic variation in *SLC6A4* influences cognitive and affective outcomes in later life. Specifically, the high-activity genotype was associated with larger right hippocampal volume in older adults, an effect that was not observed in adolescents. The gradual reduction of various brain regions including hippocampus with aging is consistently observed (51,52). Therefore, this positive effect should be considered that volume reduction was attenuated by the high promoter activity. Although the standardized effect size of the high-activity genotype on hippocampal volume is modest, this magnitude is consistent with expectations for common regulatory variants influencing complex brain traits in population-based samples. Importantly, the association remained significant after adjustment for age and sex and was robust to both covariate-based and ICV-normalized analyses (53,54), supporting an ICV-independent and biologically meaningful effect on hippocampal structure.

Subfield analyses further refined this association, implicating the hippocampal tail and the GC.ML.DG body as regions particularly sensitive to promoter activity. Brain imaging studies suggested that these regions play important roles in major depressive disorder (MDD) and cognitive decline. Their volumes were reported to be associated with responsiveness for antidepressants (55–57) and cognitive decline (58–60). The reduced volumes of the GC.ML.DG in MDD (57) and the hippocampal tail and GC.ML.DG body in subclinical geriatric depression were reported (61).

Importantly, hippocampal volume mediated the relationship between the high-activity genotype and both cognitive function and depressive tendency, whereas no direct associations between genotype and clinical scores were observed. This pattern supports an indirect pathway in which genetic variation shapes brain structure, which in turn influences clinical outcomes. Such a mechanism aligns with prior studies suggesting that the effects of serotonergic genes on behavior are often expressed through intermediate neural phenotypes rather than through direct genotype-phenotype associations (4,62,63).

The lateralization of effects to the right hippocampus warrants consideration. Although bilateral trends were observed (**Supplementary Table 8**), statistically significant associations were restricted to the right hemisphere. Right hippocampal structure has been preferentially implicated in emotional memory, stress regulation, and affective processing, which may be particularly relevant in late-life depression and cognitive–affective coupling (7,14,15). Alternatively, this asymmetry may reflect differential vulnerability or compensatory processes across hemispheres during aging (64,65).

Notably, DNA methylation at *SLC6A4* did not show direct associations with hippocampal volume in either older adults or adolescents, suggesting that epigenetic modulation primarily influences functional or cognitive outcomes rather than gross brain structure. Together with the lack of genotype effects in youth, these findings suggest that the structural impact of *SLC6A4* variation on the hippocampus emerges specifically with aging.

### Comparison with previous studies on 5-HTTLPR and aging-related phenotypes

Previous studies examining the relationship between 5-HTTLPR and cognitive or affective phenotypes in older adults have yielded largely inconsistent results. Several reports, particularly in European populations, failed to detect direct associations between the conventional S/L classification and MMSE scores or hippocampal volumes, while neuroimaging studies often reported either null effects or associations that emerged only under specific contexts such as elevated stress or cortisol levels (30,32,33). Notably, in one report, associations with depressive symptoms became evident only after reclassifying low-activity L alleles (16d), as functionally equivalent to the S allele, whereas analyses based solely on the traditional binary S/L classification remained negative (66). Taken together, these findings suggest that inconsistencies in the literature may largely reflect differences in allele classification schemes, population-specific allele frequencies, and analytical strategies. Our findings also highlight the importance of population-aware analyses for interpreting genetic influences and show that population-specific allele distributions shape vulnerability and resilience in later life. Studying diverse populations is therefore critical for uncovering biological mechanisms that may be obscured in genetically homogeneous cohorts.

### Limitations and future directions

Several limitations of this study warrant consideration. First, the observational and largely cross-sectional nature of the analyses precludes causal inference regarding the relationships among genetic variation, epigenetic regulation, brain structure, and clinical outcomes. Although mediation analyses suggest indirect pathways linking *SLC6A4* promoter activity to cognitive and affective measures, these findings should be interpreted as statistical associations rather than evidence of causality. Longitudinal studies with repeated assessments of cognitive function, mood, brain structure, and molecular markers will be essential to clarify temporal ordering and causal mechanisms. Second, DNA methylation was measured in peripheral blood or saliva, which may not fully reflect epigenetic states in brain tissue. While prior studies have reported functional CpG sites within the *SLC6A4* promoter that show cross-tissue relevance (28,29,67), tissue-specific regulation remains a critical consideration. In addition, it was reported that lifestyle factors such as smoking (68) or alcohol consumption (69) were associated with DNA methylation. However, we did not correct these influences in the present analyses. Third, for using the brain imaging data, we only focused on hippocampus, as we believe that hippocampus was one of the most rationale brain regions which were critically responsible for clinical symptoms in old participants based on the serotonergic system. Future study will include analyses in other brain regions. Finally, although the present study incorporated multiple cohorts and validated key findings across independent samples, epigenetic and neuroimaging measures were available for only a subset of participants, and sample sizes for some analyses were modest. Larger longitudinal cohorts with epigenetic and harmonized neuroimaging data (70) will improve statistical power and enable more refined analyses of gene–epigenome–brain interactions across the aging process.

## Methods

### Arao cohort study

The Arao cohort study includes older people (*N* = 1,577) of 65 years old living in Arao city, Kumamoto prefecture, Japan. This cohort is a part of JPSC-AD (34). After excluding participants without consent for genetic analyses, those with relevant medical histories, and those with missing data, data from 1,317 participants were available for analysis. Medical histories, including stroke, Parkinson’s disease and other neurological disorders, were selected for exclusion based on preliminary regression analyses showing significant associations with hippocampal volume or GDS or MMSE scores. Subjects with dementia or depression were included in the initial analyses, as these conditions constituted the primary targets of the study. Available data included age, sex, MMSE (36,37) score, GDS (38–41) score, and hippocampal brain image data, 5-HTTLPR genotypes, and DNA methylation measurements. Demographic characteristics and clinical scores are summarized in **Supplementary Table 2**. 5-HTTLPR genotyping by Sanger sequencing had been performed previously (24), whereas DNA methylation analyses by pyrosequencing were conducted in the present study (**Supplementary Methods**). Brain MRI data were acquired using 1.5-Tesla scanners and analyzed for volumetric quantification according to previously published protocols (71). Detailed information on scanner models, acquisition sequences, imaging parameters, and volumetric procedures is provided in the **Supplementary Methods**.

### JPSC-AD study

The JPSC-AD study consists of eight community-dwelling cohorts in Japan (**Extended Data Fig 1a**). Demographic characteristics are summarized in **Supplementary Table 4**. We selected 7,889 participants from seven cohorts, excluding the Arao cohort, who provided consent for genetic analyses and had no medical histories as described above or missing data for age, sex, MMSE or GDS scores. Genome-wide SNP genotyping in the JPSC-AD had been conducted previously (72), and these data were used to impute 5-HTTLPR alleles accounting for their complexity (42) using PHASE software version 2.1 (73,74). Written informed consent was obtained from all participants.

### pn-TTC study

The Tokyo Teen Cohort (TTC) study is a longitudinal, population-based cohort designed to investigate adolescent development (45). The population-neuroscience study of the TTC (pn-TTC) recruited subsamples of parent–adolescent pairs from the TTC to examine neurobiological substrates of adolescent development (35). Longitudinal data were collected at ages 11, 13, 15, and 18 years, corresponding to pn-TTC-1 through pn-TTC-4. Demographic characteristics are summarized in **Supplementary Table 6**. Brain imaging data were processed as described in the **Supplementary Methods**. 5-HTTLPR genotyping was performed using saliva DNA samples via Sanger sequencing, as previously described (24). DNA methylation analyses by pyrosequencing using saliva samples in pnTTC-1 through pn-TTC-3 had been performed previously (44). Written informed consent was obtained from adolescents and their primary parents.

### Statistical analysis

Statistical analyses were performed using R version 4.5.1 (R Foundation for Statistical Computing, Vienna, Austria) and in-house scripts, or in a CentOS Linux 7.5.1804 environment. Differences in clinical scores among 5-HTTLPR genotype subgroups were assessed using the Kruskal–Wallis test or Steel’s multiple comparison test implemented. Associations between MMSE and GDS scores were evaluated using Pearson correlation analysis. Differences in DNA methylation levels and brain regional volumes were examined using the Mann-Whitney *U* test. Mediation analyses were conducted using the *mediation* package (75) to assess indirect effects of aging on clinical scores via CpG3 DNA methylation, with sex included as a covariate, and indirect effects of the 5-HTTLPR high-activity genotype on clinical scores via hippocampal volume, adjusting for age, sex, CpG3 methylation levels, ICV, and major 5-HTTLPR genotypes. Other multivariate analyses were performed using standardized multiple regression models. To evaluate the assumptions of multiple regression models, we generated quantile-quantile (Q-Q) plots of the residuals (**Supplementary Fig. 1**). Meta-analyses across seven JPSC-AD cohorts were conducted using the *metafor* package (76). Analyses based on linear mixed models were conducted using the *lme4* (77) and *lmerTest* (78) packages. Scripts are available upon request.

## Ethics

The present study was conducted in accordance with the Declaration of Helsinki and was approved through a centralized ethics review by the Kyushu University Institutional Board for Clinical Research (Approval No. 24116). Additional site-specific approvals for study conduct were also obtained from each participating institution (Approval No. genome 341 and 529). Written informed consent was obtained from all participants.

## Data availability

Data is provided within the manuscript or supplementary information files.

## Code availability

The scripts used in the present study are available from corresponding authors on request.

## Acknowledgements

We would like to thank the participants for the contribution of their time to the JPSC-AD study and TokyoTEEN cohort. We would like to gratefully acknowledge the diligent work and contributions of all researchers and investigators in the JPSC-AD Study Group ant Tokyo TEEN cohort.

## Author contributions

YY, MB, and KI conceptualized the study. YY, NK, S. Koike, T. Ikegame, TN, and Y. Taki curated the data. YY, IN, JO, NK, S. Kikkawa, S. Koike, T. Ikegame, and YN performed the formal analysis. YY, AF, EK, KY, NK, NS, RW, S. Kikkawa, S. Koike, T. Ikegame, TN, TO, YF, YM, and MB conducted the investigation. YY, IN, JO, S. Kikkawa, S. Koike, T. Ikegame, and YN developed the methodology. KK, MT, NK, S. Koike, TN, MB, and KI administered the project. BT, HS, KK, KY, MH, MM, MT, NK, NM, NO, T. Ishikawa, TN, TO, Y. Takano, Y. Taki, Y. Tatewaki, YF, YH, YN, MB, and KI provided the resources. YN, MB, and KI supervised the study. YY performed the visualization. YY and KI wrote the original draft. All authors reviewed and edited the manuscript.

## Extended Data

**Extended Data Fig. 1.**
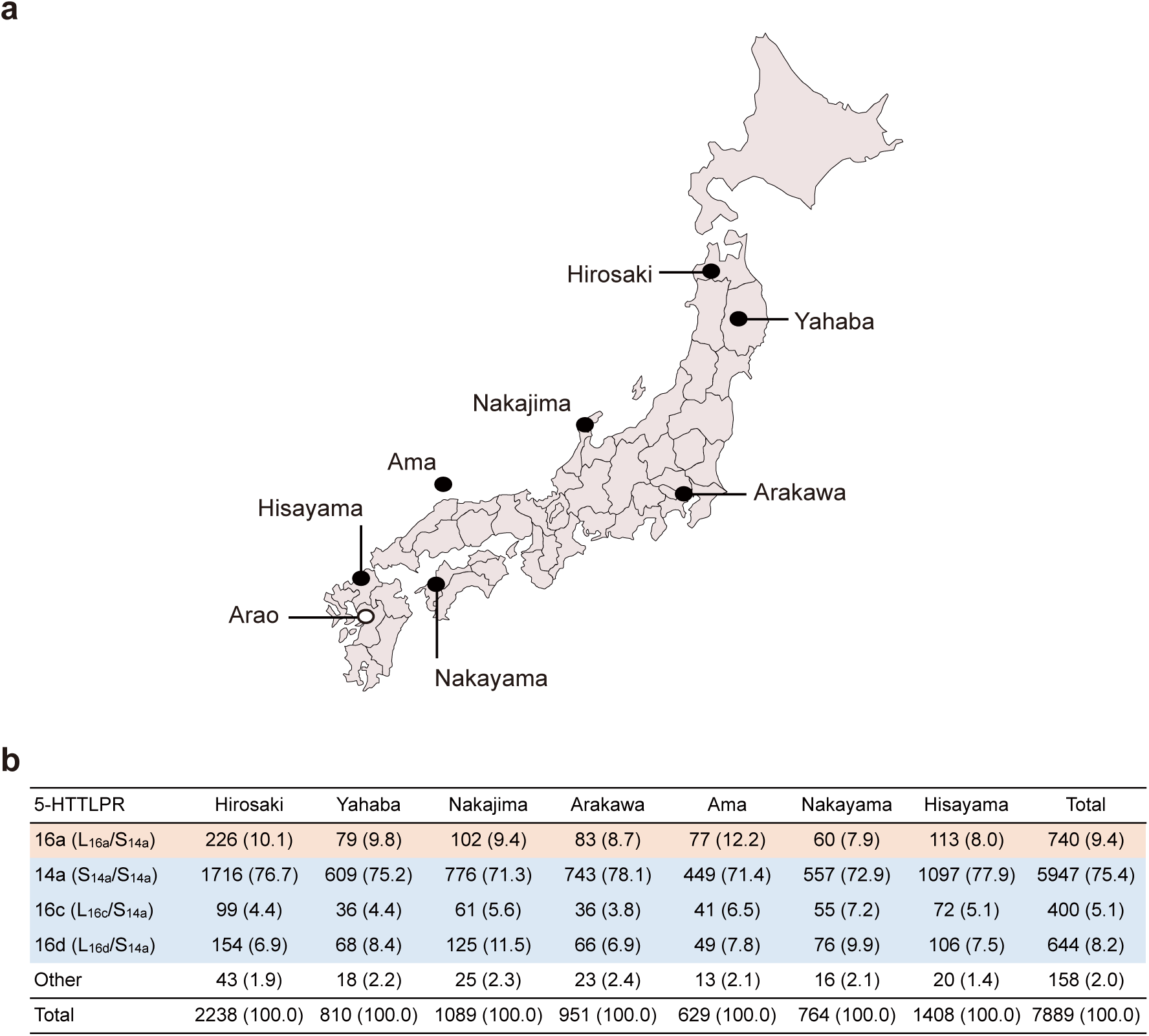
Imputed frequencies of major 5-HTTLPR genotypes across JPSC-AD cohorts. **a,** Map showing the locations of the Arao cohort and seven additional population-based cohorts included in the JPSC-AD study. Cohorts are geographically distributed across Japan, supporting the nationwide representativeness and robustness of the analyses. **b,** The number and proportion (%, in parentheses) of participants with each 5-HTTLPR genotype, estimated by imputation, across the seven JPSC-AD cohorts. Subjects were classified into four major genotypes following the Arao cohort, with 16a denoting the high-activity genotype and 14a, 16c, and 16d denoting low-activity genotypes.

**Extended Data Fig. 2.**
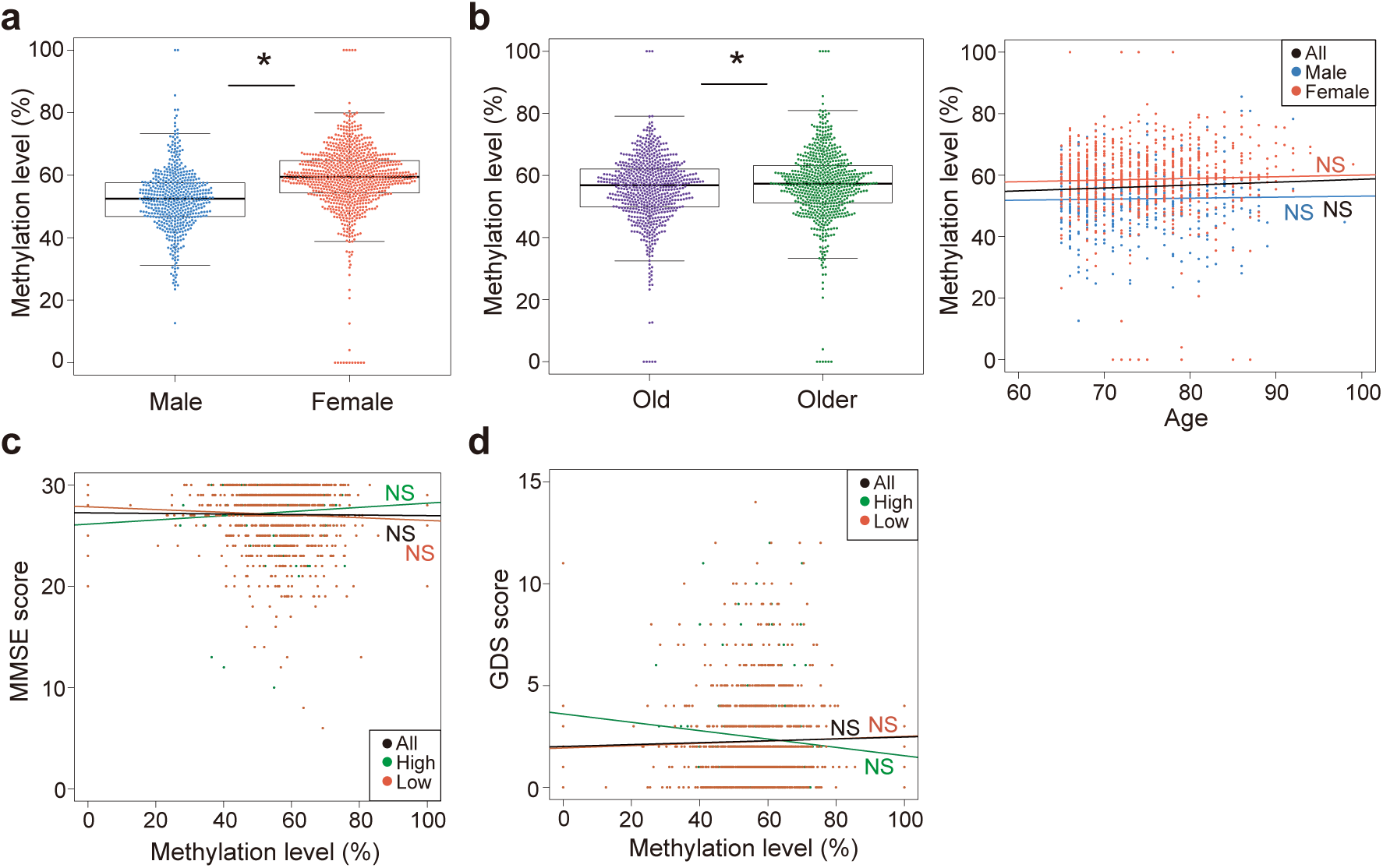
CpG4 methylation associated with sex, aging, and clinical scores. **a,** DNA methylation levels at CpG4 in males (*N* = 507) and females (*N* = 810). **b,** DNA methylation levels at CpG4 increased with age. (*left*) Methylation levels at CpG4 in old (*N* = 671) and older (*N* = 646) participants, divided by the median age of 74. (*right*) Correlation analysis with age. Black, blue, and red colors indicate individual data or regression line of all participants, males, and females, respectively. **c,** Correlation analysis with the MMSE score. Black, green, and brown colors indicate individual data or regression line of all participants, participants with the high-activity genotype (*N* = 154), and those with low-activity genotypes (*N* = 1,054), respectively. **d,** Correlation analysis with the GDS score. Box plots indicate median, interquartile range, and whiskers extending to 1.5× the interquartile range. Scatter plots show individual data points with fitted regression lines. Asterisks indicate *P*-value < 0.05. NS, not significant.

**Extended Data Fig. 3.**
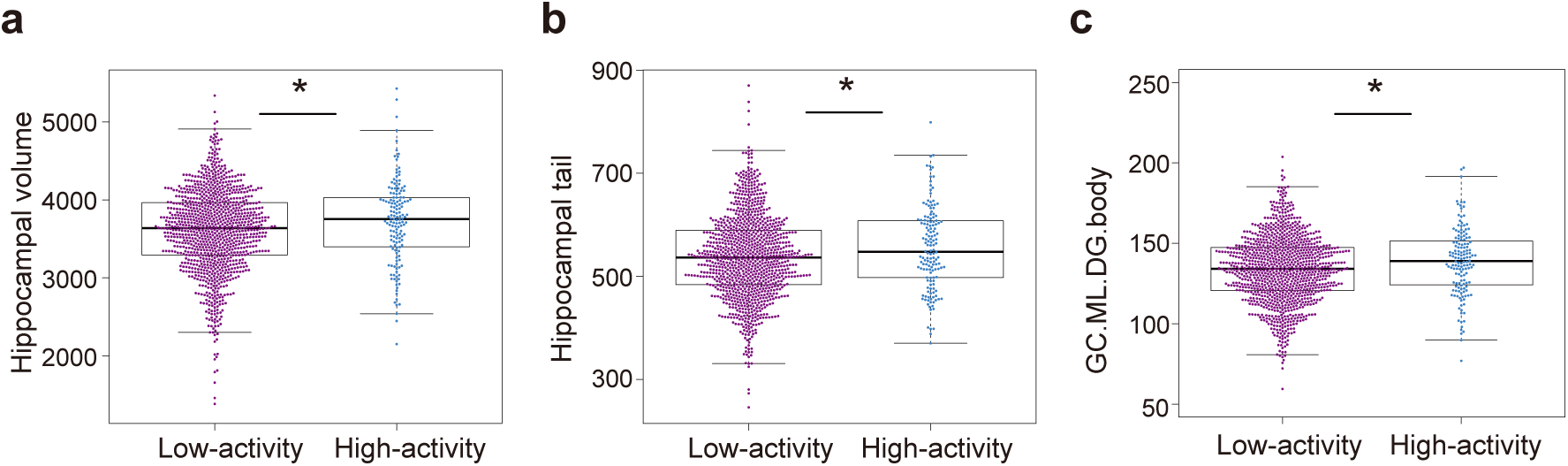
Directionally consistent differences in right hippocampal volumes by 5-HTTLPR promoter activity. Comparison of the right hippocampal volumes of the whole hippocampus (**a**), hippocampal tail (**b**), and GC.ML.DG.body (**c**). Subjects were grouped by promoter activity. Each point represents an individual participant, and boxplots show the median and interquartile range. The vertical axis indicates brain volume (mm3). Asterisks indicate *P*-value < 0.05 in Mann-Whitney *U* test.

**Extended Data Fig. 4.**
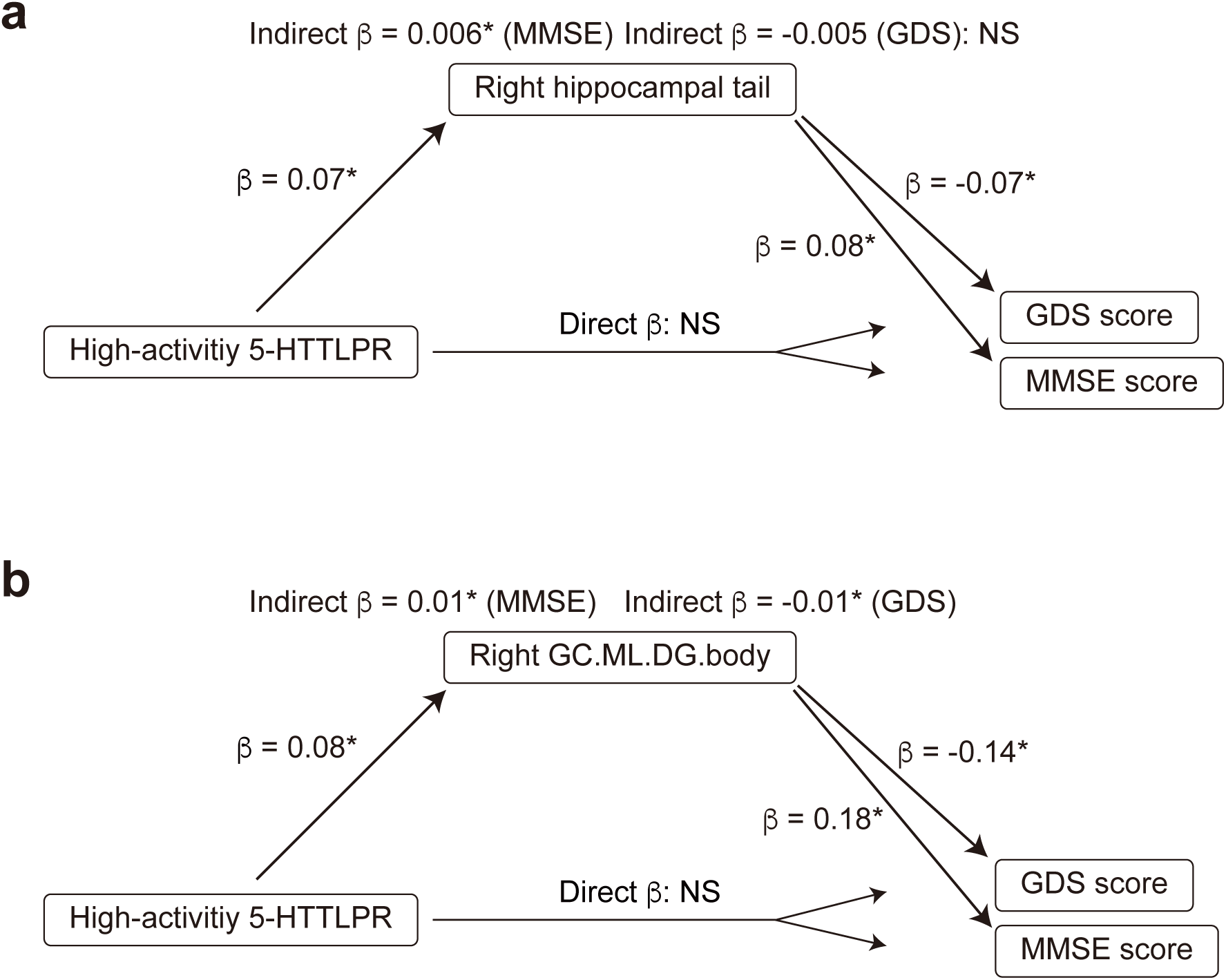
Mediation analysis via volumes of right hippocampal subfields. Mediation models illustrating the relationships among the high-activity 5-HTTLPR genotype, volumes of right hippocampal tail (**a**) and GC.ML.DG.body (**b**), and clinical outcomes. Path coefficients (β) are shown for each association. The high-activity genotype was positively associated with right hippocampal volume, which in turn was significantly associated with MMSE and GDS scores. No significant direct effects of the high-activity genotype on MMSE or GDS scores were observed. In contrast, in most cases, significant indirect effects mediated by volumes of right hippocampal tail and GC.ML.DG.body were detected for both MMSE and GDS, indicating that these structures mediate the association between 5-HTTLPR promoter activity and cognitive and depressive measures. Asterisks indicate *P*-value < 0.05. NS, not significant.

**Extended Data Table 1.** Multiple regression analysis of the right hippocampal subfield volumes. Results of a standardized multiple regression analysis examining the factors associated with right hippocampal tail and GC.ML.DG.body in older adults (*N* = 1,267). ICV, intracranial volume; SE, standard error; Statistic, *t*-statistic; CI, confidence interval.

| **Hippocampal tail** | **$\beta$** | **SE** | **Statistic** | ***P*-value** | **95% CI** |
| --- | --- | --- | --- | --- | --- |
| Age | -0.39 | 0.02 | -15.71 | < 0.01 | -0.44, -0.34 |
| Sex | -0.02 | 0.03 | -0.49 | 0.63 | -0.08, 0.05 |
| ICV | 0.25 | 0.03 | 7.80 | < 0.01 | 0.19, 0.31 |
| 16a (L<sub>16a</sub>/S<sub>14a</sub>) | 0.07 | 0.04 | 2.07 | 0.04 | 0.004, 0.14 |
| 14a (S<sub>14a</sub>/S<sub>14a</sub>) | 0.07 | 0.04 | 1.51 | 0.13 | -0.02, 0.15 |
| 16c (L<sub>16c</sub>/S<sub>14a</sub>) | 0.03 | 0.03 | 0.85 | 0.40 | -0.04, 0.09 |
| 16d (L<sub>16d</sub>/S<sub>14a</sub>) | 0.02 | 0.04 | 0.59 | 0.56 | -0.05, 0.09 |
| **GC.ML.DG.body** | **$\beta$** | **SE** | **Statistic** | ***P*-value** | **95% CI** |
| Age | -0.44 | 0.02 | -18.62 | < 0.01 | -0.49, -0.39 |
| Sex | 0.07 | 0.03 | 2.31 | 0.02 | 0.01, 0.13 |
| ICV | 0.33 | 0.03 | 10.82 | < 0.01 | 0.27, 0.39 |
| 16a (L<sub>16a</sub>/S<sub>14a</sub>) | 0.08 | 0.03 | 2.22 | 0.03 | 0.01, 0.14 |
| 14a (S<sub>14a</sub>/S<sub>14a</sub>) | 0.06 | 0.04 | 1.44 | 0.15 | -0.02, 0.14 |
| 16c (L<sub>16c</sub>/S<sub>14a</sub>) | 0.03 | 0.03 | 1.02 | 0.31 | -0.03, 0.09 |
| 16d (L<sub>16d</sub>/S<sub>14a</sub>) | 0.03 | 0.03 | 0.81 | 0.42 | -0.04, 0.09 |

## Supplementary Information

**Supplementaryentary Figure 1.**
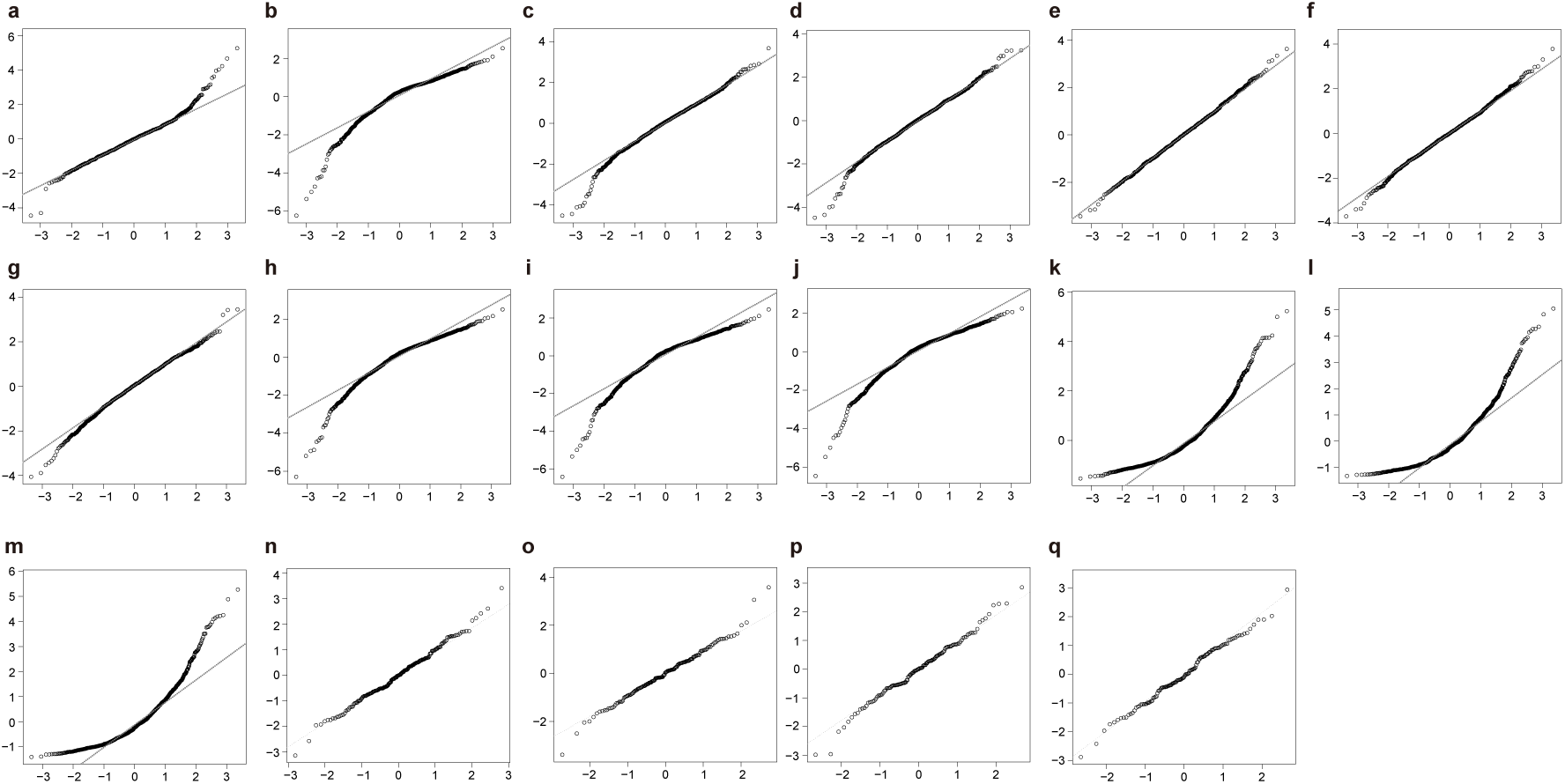
Quantile-quantile (Q-Q) plots of residuals from multiple regression analyses. Panels display Q-Q plots of residuals for models predicting the following dependent variables. In all multiple regression models, age and sex were utilized as covariates. Additional covariates for each model are shown in parentheses. The vertical and horizontal axes indicate standardized residuals and theoretical quantiles, respectively. The dashed line represents the reference line (*y* = *x*). (**a**) CpG3 methylation; (**b**) MMSE score (with CpG3 methylation); (**c**) Right hippocampal volume (with intracranial volume [ICV] and 5-HTTLPR genotype); (**d**) Ratio of right hippocampal volume to ICV (with 5-HTTLPR genotype); (**e**) Right hippocampal tail volume (with ICV and 5- HTTLPR genotype); (**f**) Right hippocampal GC.ML.DG.body volume (with ICV and 5-HTTLPR genotype); (**g**) Left hippocampal volume (with ICV and 5-HTTLPR genotype); (**h**) MMSE score (with ICV, right hippocampal volume, 5-HTTLPR genotype, and CpG3 methylation); (**i**) MMSE score (with ICV, right hippocampal tail volume, 5-HTTLPR genotype, and CpG3 methylation); (**j**) MMSE score (with ICV, right hippocampal GC.ML.DG.body volume, 5-HTTLPR genotype, and CpG3 methylation); (**k**) GDS score (with ICV, right hippocampal volume, 5-HTTLPR genotype, and CpG3 methylation); (**l**) GDS score (with ICV, right hippocampal tail volume, 5-HTTLPR genotype, and CpG3 methylation); (**m**) GDS score (with ICV, right hippocampal GC.ML.DG.body volume, 5-HTTLPR genotype, and CpG3 methylation); (**n**) ICV-standardized right hippocampal volume in pn-TTC-1 (with 5-HTTLPR genotype); (**o**) ICV-standardized right hippocampal volume in pn-TTC-2 (with 5-HTTLPR genotype); (**p**) ICV-standardized right hippocampal volume in pn-TTC-3 (with 5-HTTLPR genotype); (**q**) ICV-standardized right hippocampal volume in pn-TTC-4 (with 5-HTTLPR genotype).

**Supplementary Table 1.** Allele frequency of 5-HTTLPR in the Arao cohor. Genotyping of 5-HTTLPR was previously conducted (Ikegame et al., 2021). Major four alleles were mainly analyzed in this study.

| Allele | N | Frequency (%) |
| --- | --- | --- |
| S<sub>14a</sub> | 2073 | 78.7 |
| L<sub>16a</sub> | 196 | 7.4 |
| L<sub>16c</sub> | 125 | 4.7 |
| L<sub>16d</sub> | 179 | 6.8 |
| Other | 61 | 2.3 |
| Total | 2634 | 100.0 |

**Supplementary Table 2.** Demographic information of the Arao cohort. Demographic and clinical characteristics of participants in the Arao cohort stratified by 5-HTTLPR genotype. Shown are the number of participants (N), proportion of males, age, cognitive function assessed by the Mini-Mental State Examination (MMSE), and depressive tendency assessed by the Geriatric Depression Scale (GDS). Age, MMSE, and GDS values are presented as mean ± standard deviation. Genotypes are defined based on detailed 5-HTTLPR allelic classification. The “Other” category includes less frequent heterogenous genotypes that were not included in the main analyses. Obvious biases were not observed (Kruskal-Wallis test, *P*-value > 0.05).

| Genotype | N | Male (%) | Age | MMSE | GDS |
| --- | --- | --- | --- | --- | --- |
| 14a (S<sub>14a</sub>/S<sub>14a</sub>) | 816 | 38.8 | 74.1 ± 6.5 | 27.0 ± 3.0 | 2.2 ± 2.2 |
| 16a (L<sub>16a</sub>/S<sub>14a</sub>) | 154 | 41.6 | 73.2 ± 6.3 | 27.3 ± 3.2 | 2.4 ± 2.6 |
| 16c (L<sub>16c</sub>/S<sub>14a</sub>) | 96 | 36.5 | 73.6 ± 6.6 | 27.6 ± 3.1 | 2.6 ± 2.5 |
| 16d (L<sub>16d</sub>/S<sub>14a</sub>) | 142 | 37.3 | 73.8 ± 5.9 | 27.3 ± 2.9 | 2.2 ± 2.1 |
| Other | 109 | 34.9 | 75.6 ± 7.0 | 26.9 ± 2.8 | 2.2 ± 2.3 |
| Total | 1317 | 38.5 | 74.0 ± 6.5 | 27.1 ± 3.0 | 2.3 ± 2.3 |

**Supplementary Table 3.** Estimated allele frequency of 5-HTTLPR in the JPSC-AD. Estimated allele frequency of 5-HTTLPR in the JPSC-AD participants (*N* = 7,889).

| Allele | N | Frequency (%) |
| --- | --- | --- |
| S<sub>14a</sub> | 13678 | 86.7 |
| L<sub>16a</sub> | 892 | 5.7 |
| L<sub>16c</sub> | 451 | 2.9 |
| L<sub>16d</sub> | 757 | 4.8 |
| Total | 15778 | 100.0 |

**Supplementary Table 4.** Demographic and clinical characteristics by imputed 5-HTTLPR genotype across JPSC-AD cohorts. Genotypes were grouped into four major classes following the Arao cohort classification. Values are shown as number of participants (N), percentage of males, and mean ± standard deviation for age, Mini-Mental State Examination (MMSE), and Geriatric Depression Scale (GDS) scores. Genotypes were inferred by imputation using a method accounting for the complexity of 5-HTTLPR variants. Obvious biases were not observed (Kruskal-Wallis test, *P*-value > 0.05).

| Genotype | N | Male (%) | Age | MMSE | GDS |
| --- | --- | --- | --- | --- | --- |
| 14a (S<sub>14a</sub>/S<sub>14a</sub>) | 5947 | 41.8 | 72.7 ± 6.6 | 27.2 ± 3.1 | 2.9 ± 2.7 |
| 16a (L<sub>16a</sub>/S<sub>14a</sub>) | 740 | 42.8 | 72.8 ± 6.9 | 27.1 ± 3.3 | 2.9 ± 2.8 |
| 16c (L<sub>16c</sub>/S<sub>14a</sub>) | 400 | 41.3 | 73.2 ± 7.0 | 27.2 ± 2.9 | 2.8 ± 2.5 |
| 16d (L<sub>16d</sub>/S<sub>14a</sub>) | 644 | 45.0 | 72.9 ± 6.8 | 27.1 ± 3.1 | 2.9 ± 2.6 |
| Other | 158 | 37.3 | 72.6 ± 7.5 | 26.9 ± 3.3 | 2.5 ± 2.5 |
| Total | 7889 | 42.1 | 72.7 ± 6.7 | 27.2 ± 3.1 | 2.9 ± 2.7 |

**Supplementary Table 5.** Multiple regression analysis of ICV-normalized right hippocampal volume. Results of a standardized multiple regression analysis using the proportion of the right hippocampal volume relative to intracranial volume (ICV-normalized hippocampal volume) (*N* = 1,286). SE, standard error; Statistic, *t*-statistic; CI, confidence interval.

| Variable | $\beta$ | SE | Statistic | *P*-value | 95% CI |
| --- | --- | --- | --- | --- | --- |
| Age | -0.46 | 0.02 | -19.58 | < 0.01 | -0.51, -0.42 |
| Sex | 0.33 | 0.02 | 13.99 | < 0.01 | 0.28, 0.38 |
| 16a (L<sub>16a</sub>/S<sub>14a</sub>) | 0.07 | 0.03 | 2.00 | 0.045 | 0.001, 0.14 |
| 14a (S<sub>14a</sub>/S<sub>14a</sub>) | 0.07 | 0.04 | 1.62 | 0.11 | -0.01, 0.15 |
| 16c (L<sub>16c</sub>/S<sub>14a</sub>) | 0.05 | 0.03 | 1.49 | 0.14 | -0.01, 0.11 |
| 16d (L<sub>16d</sub>/S<sub>14a</sub>) | 0.04 | 0.03 | 1.24 | 0.21 | -0.02, 0.11 |

**Supplementary Table 6.** Demographic information of the pn-TTC. Demographic characteristics of the pn-TTC cohort across four waves: pn-TTC-1 (*N* = 202), pn-TTC-2 (*N* = 158), pn-TTC-3 (*N* = 131), and pn-TTC-4 (*N* = 123). Values are presented as the number of participants, proportion of males (%), and mean age at MRI scan with standard deviation.

|  | pn-TTC-1 |  |  | pn-TTC-2 |  |  | pn-TTC-3 |  |  | pn-TTC-4 |  |  |
| --- | --- | --- | --- | --- | --- | --- | --- | --- | --- | --- | --- | --- |
| Genotype | N | Male | Age | N | Male | Age | N | Male | Age | N | Male | Age |
| 14a (S<sub>14a</sub>/S<sub>14a</sub>) | 123 | 53.7 | 11.6 ± 0.7 | 98 | 53.1 | 13.6 ± 0.6 | 83 | 53 | 15.8 ± 0.8 | 75 | 49.3 | 18.2 ± 0.9 |
| 16a (L<sub>16a</sub>/S<sub>14a</sub>) | 29 | 51.7 | 11.7 ± 0.8 | 21 | 47.6 | 13.6 ± 0.6 | 18 | 50 | 16 ± 0.7 | 17 | 52.9 | 18.2 ± 0.7 |
| 16c (L<sub>16c</sub>/S<sub>14a</sub>) | 12 | 33.3 | 11.3 ± 0.3 | 10 | 30 | 13.7 ± 0.9 | 7 | 14.3 | 16 ± 1.1 | 7 | 14.3 | 18 ± 0.9 |
| 16d (L<sub>16d</sub>/S<sub>14a</sub>) | 24 | 45.8 | 11.5 ± 0.8 | 17 | 52.9 | 13.4 ± 0.7 | 13 | 46.2 | 15.5 ± 0.8 | 13 | 53.8 | 17.8 ± 0.8 |
| Other | 14 | 64.3 | 11.6 ± 0.7 | 12 | 66.7 | 13.5 ± 0.4 | 10 | 60 | 15.7 ± 0.7 | 11 | 54.5 | 18.2 ± 0.9 |
| Total | 202 | 52 | 11.6 ± 0.7 | 158 | 51.9 | 13.6 ± 0.6 | 131 | 50.4 | 15.8 ± 0.8 | 123 | 48.8 | 18.1 ± 0.9 |

**Supplementary Table 7.** Multiple regression analysis of right hippocampal volumes in youth. Detailed results of multiple regression analyses using ICV-adjusted right hippocampal volume as the dependent variable (pn-TTC-1, *N* = 202; pn-TTC-2, *N* = 158; pn-TTC-3, *N* = 131; pn-TTC-4, *N* = 123). SE, standard error; Statistic, *t*-statistic; CI, confidence interval.

| **pn-TTC-1** | **$\beta$** | **SE** | **Statistic** | ***P*-value** | **95% CI** |
| --- | --- | --- | --- | --- | --- |
| Age | 0.08 | 0.07 | 1.16 | 0.25 | -0.05, 0.21 |
| Sex | -0.38 | 0.07 | -5.73 | < 0.01 | -0.51, -0.25 |
| 16a (L<sub>16a</sub>/S<sub>14a</sub>) | -0.06 | 0.11 | -0.52 | 0.60 | -0.27, 0.15 |
| 14a (S<sub>14a</sub>/S<sub>14a</sub>) | 0.01 | 0.13 | 0.11 | 0.91 | -0.24, 0.27 |
| 16c (L<sub>16c</sub>/S<sub>14a</sub>) | 0.002 | 0.09 | 0.02 | 0.99 | -0.17, 0.17 |
| 16d (L<sub>16d</sub>/S<sub>14a</sub>) | 0.09 | 0.10 | 0.90 | 0.37 | -0.11, 0.29 |
| **pn-TTC-2** | **$\beta$** | **SE** | **Statistic** | ***P*-value** | **95% CI** |
| Age | -0.04 | 0.07 | -0.58 | 0.56 | -0.18, 0.10 |
| Sex | -0.45 | 0.07 | -6.22 | < 0.01 | -0.59, -0.31 |
| 16a (L<sub>16a</sub>/S<sub>14a</sub>) | 0.02 | 0.11 | 0.16 | 0.87 | -0.20, 0.24 |
| 14a (S<sub>14a</sub>/S<sub>14a</sub>) | 0.14 | 0.13 | 1.01 | 0.31 | -0.13, 0.40 |
| 16c (L<sub>16c</sub>/S<sub>14a</sub>) | -0.04 | 0.09 | -0.41 | 0.68 | -0.23, 0.15 |
| 16d (L<sub>16d</sub>/S<sub>14a</sub>) | 0.13 | 0.10 | 1.21 | 0.23 | -0.08, 0.33 |
| **pn-TTC-3** | **$\beta$** | **SE** | **Statistic** | ***P*-value** | **95% CI** |
| Age | -0.07 | 0.08 | -0.88 | 0.38 | -0.22, 0.08 |
| Sex | -0.54 | 0.08 | -7.07 | < 0.01 | -0.69, -0.39 |
| 16a (L<sub>16a</sub>/S<sub>14a</sub>) | 0.01 | 0.12 | 0.12 | 0.90 | -0.22, 0.25 |
| 14a (S<sub>14a</sub>/S<sub>14a</sub>) | 0.003 | 0.14 | 0.02 | 0.98 | -0.27, 0.28 |
| 16c (L<sub>16c</sub>/S<sub>14a</sub>) | -0.03 | 0.10 | -0.30 | 0.76 | -0.22, 0.16 |
| 16d (L<sub>16d</sub>/S<sub>14a</sub>) | 0.02 | 0.11 | 0.20 | 0.84 | -0.19, 0.24 |
| **pn-TTC-4** | **$\beta$** | **SE** | **Statistic** | ***P*-value** | **95% CI** |
| Age | -0.10 | 0.08 | -1.31 | 0.19 | -0.25, 0.05 |
| Sex | -0.56 | 0.08 | -7.34 | < 0.01 | -0.71, -0.41 |
| 16a (L<sub>16a</sub>/S<sub>14a</sub>) | 0.01 | 0.11 | 0.10 | 0.92 | -0.21, 0.23 |
| 14a (S<sub>14a</sub>/S<sub>14a</sub>) | 0.04 | 0.13 | 0.31 | 0.76 | -0.22, 0.30 |
| 16c (L<sub>16c</sub>/S<sub>14a</sub>) | -0.02 | 0.09 | -0.21 | 0.83 | -0.21, 0.17 |
| 16d (L<sub>16d</sub>/S<sub>14a</sub>) | 0.07 | 0.11 | 0.70 | 0.49 | -0.14, 0.28 |

**Supplementary Table 8.** Multiple regression analysis of the left hippocampal volume. Results of a standardized multiple regression analysis examining the factors associated with left hippocampal volume in older adults (*N* = 1,286). ICV, intracranial volume; SE, standard error; Statistic, *t*-statistic; CI, confidence interval.

| Variable | $\beta$ | SE | Statistic | *P*-value | 95% CI |
| --- | --- | --- | --- | --- | --- |
| Age | -0.49 | 0.02 | -21.45 | < 0.01 | -0.54, -0.45 |
| Sex | 0.07 | 0.03 | 2.32 | 0.02 | 0.01, 0.13 |
| ICV | 0.30 | 0.03 | 10.14 | < 0.01 | 0.24, 0.36 |
| 16a (L<sub>16a</sub>/S<sub>14a</sub>) | 0.06 | 0.03 | 1.79 | 0.07 | -0.01, 0.12 |
| 14a (S<sub>14a</sub>/S<sub>14a</sub>) | 0.04 | 0.04 | 0.95 | 0.34 | -0.04, 0.12 |
| 16c (L<sub>16c</sub>/S<sub>14a</sub>) | 0.02 | 0.03 | 0.66 | 0.51 | -0.04, 0.08 |
| 16d (L<sub>16d</sub>/S<sub>14a</sub>) | 0.03 | 0.03 | 0.85 | 0.39 | -0.04, 0.09 |

## Supplementary Methods

### DNA methylation assay in the Arao cohort

Genomic DNA was extracted from peripheral blood cells of Arao cohort participants using the QIAamp DNA Blood Mini QIAcube Kit (QIAGEN, Hilden, Germany). Extracted DNA was treated with sodium bisulfite conversion using the EpiTect 96 Bisulfite Kit (QIAGEN). The CpG island shore near the exon 1 of *SLC6A4* (hg19 coordinates, chr17:30,235,139-30,235,342) was amplified by PCR using bisulfite-converted genomic DNA as a template. The forward primer sequence was 5′-TTTTTAGTTGTTTGGGTATTTGTGTTA-3′, and the reverse primer was 5′-biotinylated (Bio-5′-AAAACTTTACAACCTCTTAAAAACCC-3′). PCR amplification was conducted in a total reaction volume of 50 μL containing 10 μL of 5 M betaine, 5 μL of 10 × PCR buffer (Invitrogen, Carlsbad, CA, USA), 3 μL of 50 mM MgCl2, 1 μL of 10 mM each dNTP, 2 μL of 10 μM each primer, 2 ng of single-strand DNA binding (Promega, Madison, WI, USA), 1 μL of Thermo Platinum Taq (Invitrogen), 2 μL of template DNA, and double distilled water. PCR was carried out with an initial denaturation at 95 °C for 3 min, followed by 40 cycles of 98 °C for 10 s, 57 °C for 30 s, and 72 °C for 30 s. Each PCR product (2 μL) was used for microchip electrophoresis by MultiNA (Shimadzu, Kyoto, Japan) and 23 μL was subjected to pyrosequencing analysis to quantify DNA methylation levels at CpG3 and CpG4. Biotinylated PCR products were immobilized on streptavidin-coated Sepharose beads (Cytiva, Marlborough, MA, USA) and processed to generate single-stranded DNA using a PyroMark Q96 Vacuum Workstation (QIAGEN). The immobilized DNA was sequentially washed with double-distilled water, 70% ethanol, denaturation solution (0.2 N NaOH), and washing buffer (10 mM Tris-acetate, pH 7.6). The captured single-stranded DNA was released into a PSQ96 Plate (QIAGEN) containing 48 μL of PyroMark Annealing Buffer (QIAGEN), 1 μL of sequencing primer (5′-AATATAAATTATGGGTTGAA-3′; 10 μM), and 2 ng of single-strand DNA binding protein (Promega). The mixture was heated at 90 °C for 3 minutes and allowed to anneal at room temperature. Pyrosequencing was performed using PyroMark Gold Q96 reagents on a PSQ96MA instrument (QIAGEN) according to the manufacturer’s protocol. DNA methylation levels at each CpG site were calculated using PSQ96MA software (QIAGEN).

### Brain imaging analysis in the Arao cohort

Brain MRI data were acquired following a previously published protocol (Yoshiura et al., 2022). MRI scans were performed at Arao Municipal Hospital using a 1.5-Tesla Philips Ingenia CX Dual scanner (Philips Healthcare, Amsterdam, Netherlands) and at Omuta Tenryo Hospital using a 1.5-Tesla GE Signa HDxt Ver.23 scanner (GE Healthcare, Chicago, IL, USA). Potential scanner effects were assessed by including scanner type as a covariate in the regression analyses, and no significant influence of scanner differences on the results was observed.

At Arao Municipal Hospital, the Philips MRI protocol included a high-resolution three-dimensional (3D) T1-weighted sequence (repetition time [TR] = 8.6 ms, echo time [TE] = 4.0 ms, flip angle = 9°, matrix = 192 × 192, slice thickness = 1.2 mm). At Omuta Tenryo Hospital, the GE MRI protocol included a high-resolution 3D T1-weighted sequence (TR = 8.3 ms, TE = 3.4 ms, flip angle = 8°, matrix = 192 × 192, slice thickness = 1.2 mm).

Volumetric quantification was performed based on the method described previously (Shima et al., 2024) using FreeSurfer version 7.0 to analyze 3D T1-weighted images and to obtain the left and right whole hippocampal volumes and the volumes of 12 hippocampal subfields (HATA, fimbria, hippocampal fissure, molecular layer, GC-ML-DG, CA1, CA3, CA4, subiculum, presubiculum, parasubiculum, and hippocampal tail).

### Brain imaging analysis in the pn-TTC

In pn-TTC study, three acquisition procedures were used: Procedure 1 used a Philips Achieva with an 8-channel head coil and obtained 518 scans in pn-TTC-1 and pn-TTC-2. Procedure 2 used Siemens Prisma with a 64-channel head coil and obtained 125 scans in pn-TTC-2. Procedure 3 used Siemens Prisma with a 32-channel head coil and obtained 693 scans in pn-TTC-3 and pn-TTC-4.

The images using Procedures 1 and 2 were preprocessed using the legacy style in the HCP pipeline, primarily via the FreeSurfer version 6.0.1 “recon-all” command. The images obtained using Procedure 3 were preprocessed using HCP pipeline version 4.3.0 including readout distortion correction (J. Jovicich et al., 2006 and Glasser et al., 2013). The pipeline requires both T1w and T2w images and provides greater validity and reliability of cortical thickness, surface area, and subcortical volume. If T2w images were unavailable for any reason, we preprocessed only T1w images using FreeSurfer “recon-all” for Procedures 1 and 2. Since we obtained brain features using different scan procedures and image preprocessing, we applied traveling subject harmonization to the features (Yamashita et al., 2019 and Maikusa et al., 2021).

